# LOW-COST PELVIC PHANTOM PROTOTYPE WITH MATERIALS RESEMBLING HUMAN TISSUES

**DOI:** 10.1101/2024.05.23.24307760

**Authors:** Javiera García Coya, Catalina Nazar Meier, TM Marco Jiménez Herrera, Castillo V. TM Ricardo

## Abstract

**Introduction:** Phantoms are elements used in radiological services for quality control tests. In the case of equipment with ionizing radiation, quality control tests focus on image quality and dosimetry. These phantoms are made with materials that resemble human tissues internally but are high-cost and difficult to access for radiology services.

**Objectives:** To design a methodology for creating an anthropomorphic radiology phantom with materials correlating to human tissues of the pelvis for various uses, using Hounsfield Units (HU) as a comparison.

**Methodology:** Data from 34 female patients with bone pelvis exams were collected. HU was measured in bone, muscle, and adipose tissue of the hip. Simultaneously, HU of different material mixtures was measured to correlate data between patients and materials for the creation of a right hip phantom.

**Results:** HU values of the materials match human tissues except for adipose tissue, which is slightly increased. The costs for phantom creation were low.

**Discussion and Conclusion:** HU estimates of materials and human tissue mostly align with those estimated in the sample and reported in the literature, achieving the creation of a low-cost phantom easily accessible for institutions or educational centers requiring quality, dosimetry, or educational tests.

## BACKGROUND

Phantoms, also known as mannequins, are widely used devices in the healthcare field for various purposes, including education and equipment calibration, especially in radiology, to ensure image quality and dosimetry without risking animals or humans (1–4).

There are anthropomorphic phantoms made with materials of similar density to human tissues and with realistic anatomical shapes (3,4). To achieve similarity to human tissues, one must consider the linear attenuation coefficient of materials and, for radiology, the electron density, mass attenuation coefficients, and mass-energy absorption coefficients (5). The precision of these materials resembling human tissues depends on their intended purpose (5).

The relative difference of the linear attenuation coefficient compared to water is termed the Computed Tomography (CT) number, measured in Hounsfield Units (HU). This is presented under a specific voltage to characterize the tissue (4). The issue with identifying tissue using this method is that it’s specific to a particular energy(6). The Hounsfield Unit (HU) values for human tissues have been extensively studied, with muscular tissue ranging from 40 to 44 according to White et al. 1989 (5), 60 (±) 30 according to Sirtoli et al. 2017 (7) y 54 (±) 7 according to Niebuhr et al 2019(8); For cancellous bone tissue according to Mohammed et al., 2019, of 262(9) and 265 according to Sirtoli et al., 2017(7); For cortical bone tissue, 1454 according to Abdullah et al., 2018(10) and 819 (±) 211 according to Niebuhr et al., 2016(11); For adipose tissue, from -95 to -55 according to Woodard & White, 1986.(12) and Niebuhr et al., 2016(11).

For the creation of these phantoms, materials that mimic soft tissues such as muscle and compact tissues such as bone should be sought, and they should also correlate with the HU of human tissues. Regarding soft tissues, these can be solid polymethyl methacrylate (PMMA) and resins.(13)(14) and also 3D printed(15). The liquids mixed with contrast media have simulated a variety of organs and tissues (8,10,11). Furthermore, it has been demonstrated that gelatin materials exhibit similarities to soft tissues (10,16). The issue with these compounds is that radiological changes over time have been observed, but methods have been developed to maintain their state (17). Regarding bone materials, metallic materials infused in 3D printing filaments with excessively high Hounsfield Units (HU) have been utilized, resulting in artifacts in the CT image (18), on the other hand, acrylonitrile butadiene styrene (ABS) granules have yielded favorable results (19). Gypsum has also been used (8,11), And iodine solution has been used as part of the bone cortex (10).

Given that the prices of commercial phantoms are prohibitive, the need to create low-cost alternatives has been recognized (20), the purpose of this work is to create a low-cost adult pelvis phantom that accurately reproduces the characteristics of the human body, aiming to be accessible to a variety of medical centers, hospitals, and clinics. This initiative significantly contributes to quality control of equipment and precise measurement of radiological dosage by seeking materials that resemble human tissues through Hounsfield Units (HU). Additionally, it will serve as a valuable pedagogical tool for teaching anatomy and radiological positioning.

## METODOLOGY

This research has been approved by the ethics committee of our institution. The methodology consisted of three stages.

### Stage I. Extraction of human tissue data

Thirty-four computed tomography (CT) scans of pelvic bone from female patients aged between 30 and 55 years were selected, meeting the inclusion criteria, which included no history of hip pathology, fractures, hip prostheses, osteopenia, or any element that could alter the measurement of Hounsfield Units (HU). HUs were measured using a standardized 1 cm2 Region of Interest (ROI) in different anatomical zones of bone, muscle, and adipose tissues. Bone tissue involved the proximal third of the femur and the acetabulum, comprising the hip joint, and was specifically measured in the right hip. The femur was assessed for head, neck, shaft, and cortical measurements; the acetabulum for its medial, central, and lateral margins; the muscle for its proximal lateral, distal lateral, and distal medial portions; and the fat for its proximal lateral, distal lateral, and distal medial regions. These data will subsequently be compared with the HU of materials simulating human tissue in Stage II. The acetabulum measurement is illustrated in Figure 1.

**Figure 1.**
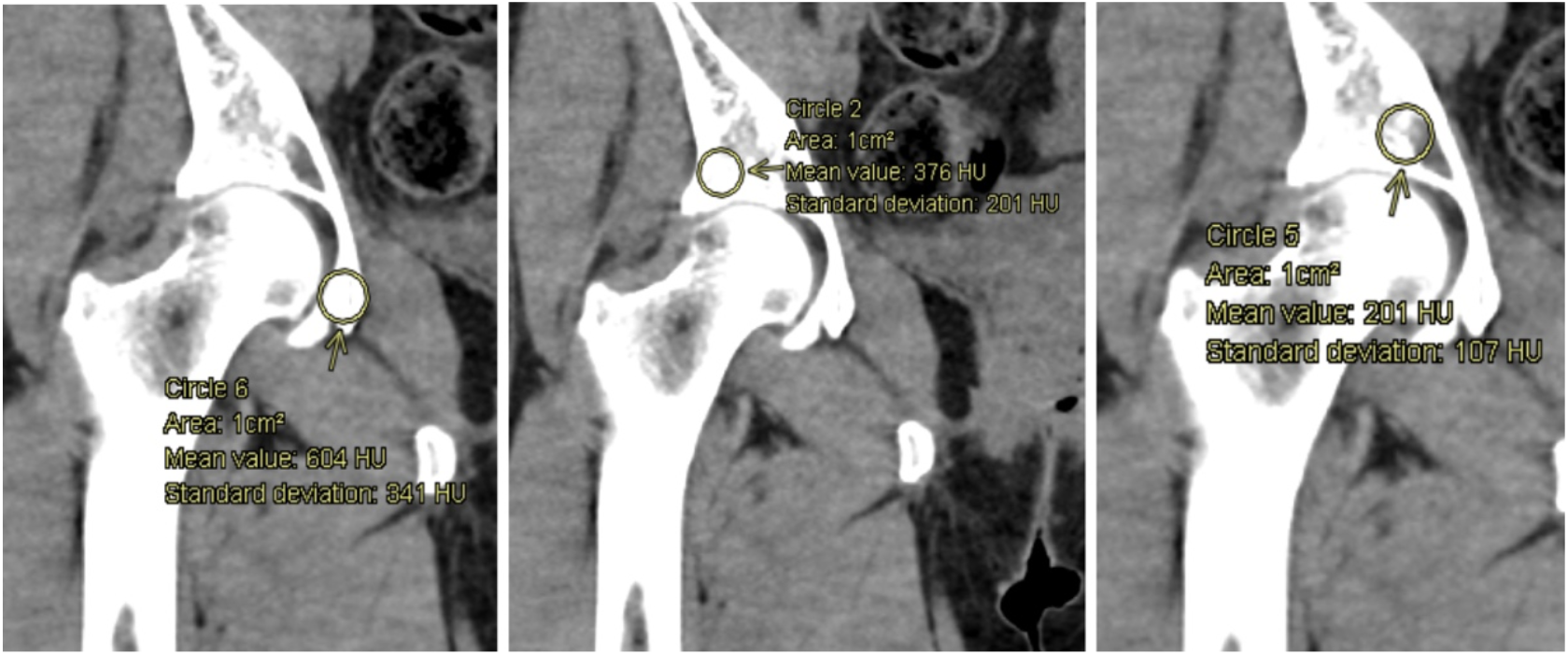
Measurement of Hounsfield Units (HU) in the acetabulum. Measurement of Hounsfield Units (HU) in different anatomical zones using a standardized 1 cm2 Area Region of Interest (ROI).

### Stage II. Material selection and Hounsfield Unit (HU) measurement

This stage consisted of 5 tests plus a final test, where a Computed Tomography (CT) scan (Figure 2) was taken of each test containing different materials and mixtures with various proportions to correlate with the density of real human tissue. The materials and mixtures used were as follows: 30% ballistic gel + 6 cubic centimeters (cc) of iodinated contrast medium (ICM), 30% ballistic gel + 3 cc of ICM, Plaster (20 g) + ultrasound gel (20 ml), 10% ballistic gel, 15% ballistic gel, Vaseline (10 g) + 10 grams (g) of flour, 30% ballistic gel + 7 cc of ICM, 30% ballistic gel + 3.5 cc of ICM. Subsequently, the Hounsfield Units (HU) of each material were measured.

**Figure 2.**
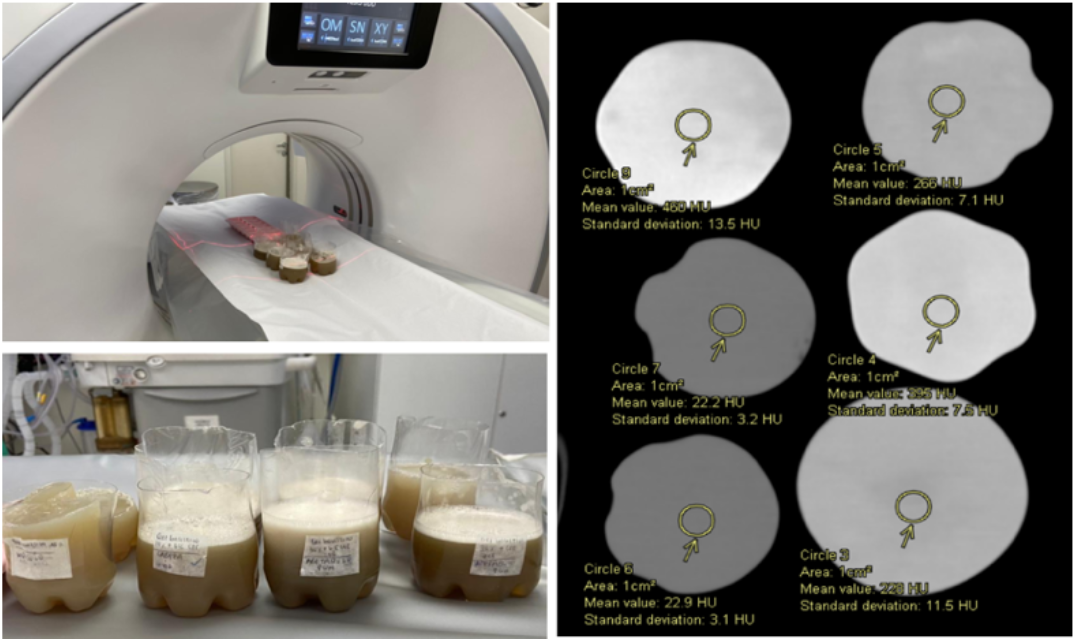
Preparation and execution of the CT scan for each mixture. Computed Tomography of the materials with their respective Hounsfield Unit (HU) measurements using ROI.

### Stage III. Phantom construction

Two mock-ups were created. The first mock-up aimed at the complete modeling of the phantom through 3D printing (Figure 3), meaning a shell of the entire phantom to be later filled with the selected materials. However, the complexity of the mold that would be created resulted in some issues with the software modeling, leading to a scale-sized phantom with a construction that couldn’t be filled with liquid materials due to holes where they could leak (Figure 3b).

**Figure 3.**
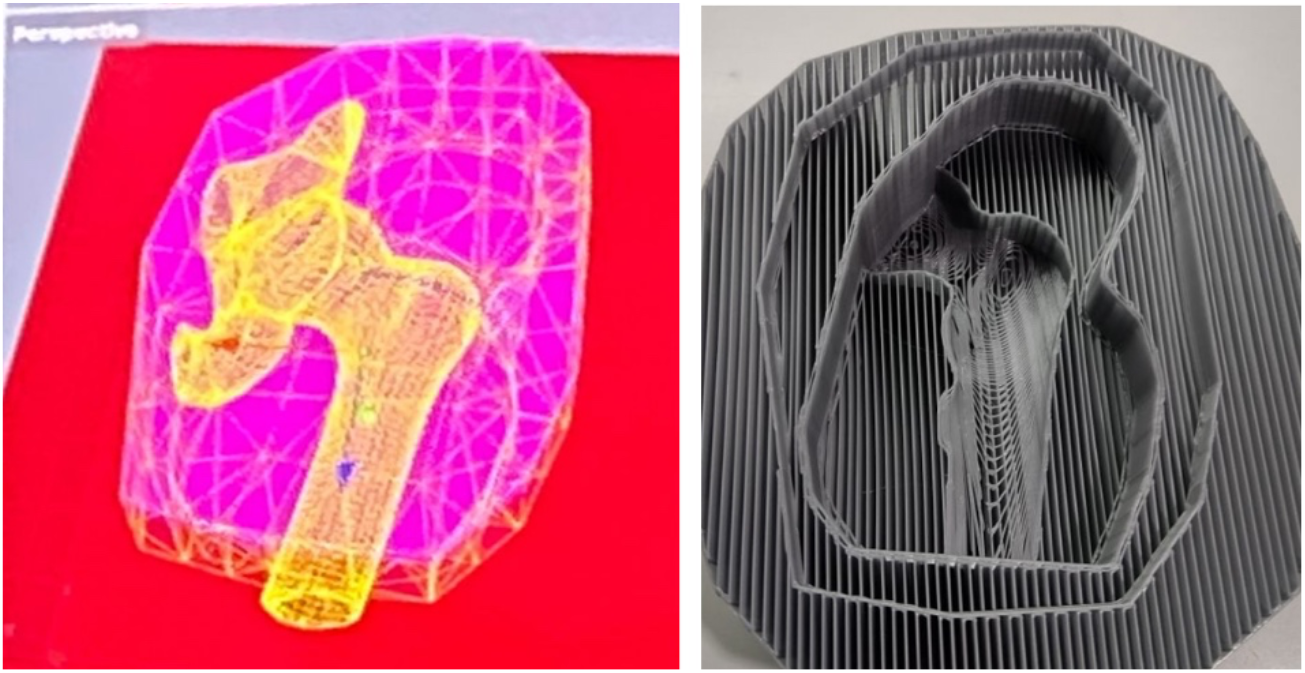
Modeling of the first mock-up with 3D printing. The modeling was carried out using Rhinoceros V7 software (left image), and the result of the mock-up is shown in the (right image)

The second mock-up was made in a completely different manner. The proximal third of the femur and the acetabulum were 3D printed in positive (Figure 4), to obtain a shape to then be placed in a rubber silicone mold. The 3D models were then impressed into the silicone rubber. Simultaneously, the selected mixtures were prepared according to the obtained densities. The mixture that resembled the density of the femur corresponded to 30% ballistic gel plus 6 cc of contrast, which was initially a liquid mixture allowing it to fill the femur mold in the silicone. It was then left to dry, demolded, and the femur shape was obtained. Subsequently, the muscle mixture corresponding to 15% ballistic gel was prepared. The 3D model of the femur and the acetabulum were placed at the bottom of a container, allowing the ballistic gel mixture to be poured over them. After drying, the femur shape formed the joint with the acetabulum (Figure 4b). The same technique was used to fill the acetabulum, which for practical reasons, was decided to be made with the same density as the femur. Finally, the adipose tissue was created using a mixture of vaseline and flour in equal proportion. The final phantom, without the fat, was placed in a plastic box for support, and on the sides, it was filled with the corresponding fat material to obtain the final phantom model (Figure 4).

**Figure 4.**
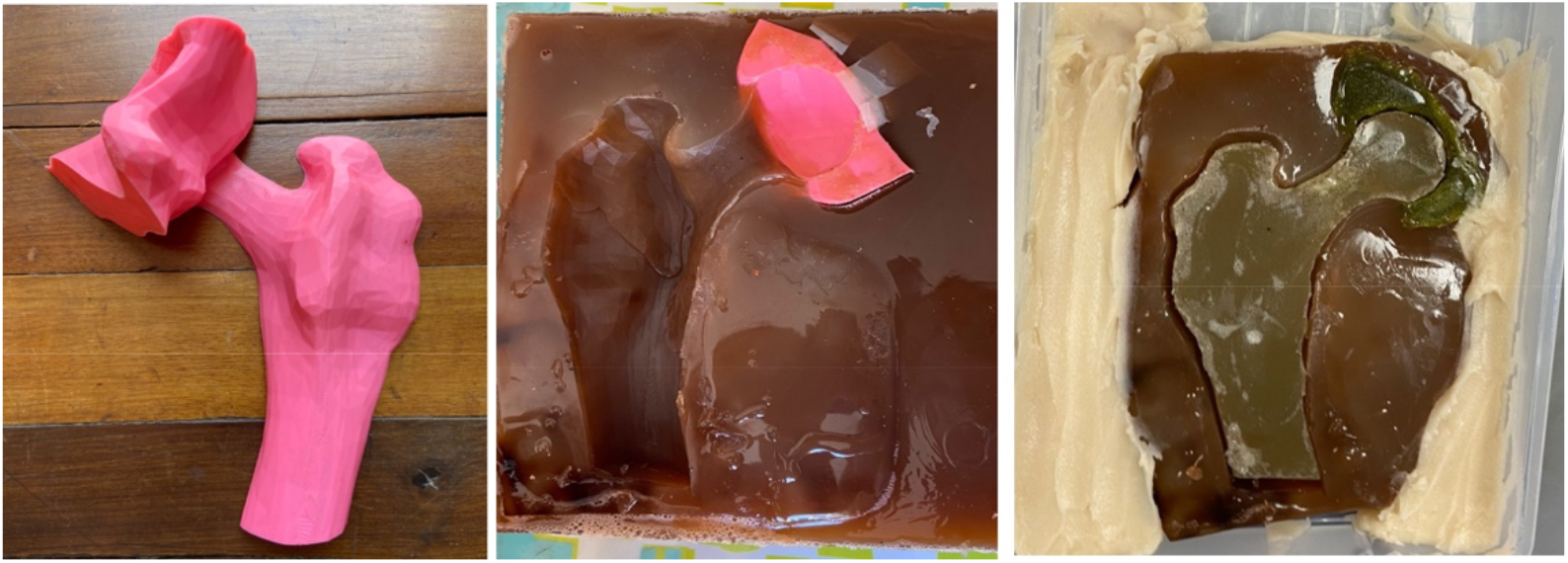
Modeling of the second mock-up. Hip joint in 3D print (left image). Shape of the femur and acetabulum in the ballistic gel mold corresponding to the muscle (center image). Final phantom with the fat mixture on the periphery (right image).

Subsequently, a Computed Tomography (CT) scan with a bony pelvis protocol was performed on the final phantom (Figure 5), and multiple Regions of Interest (ROIs) were measured across various tissues and anatomical areas that mimicked the materials.

**Figure 5.**
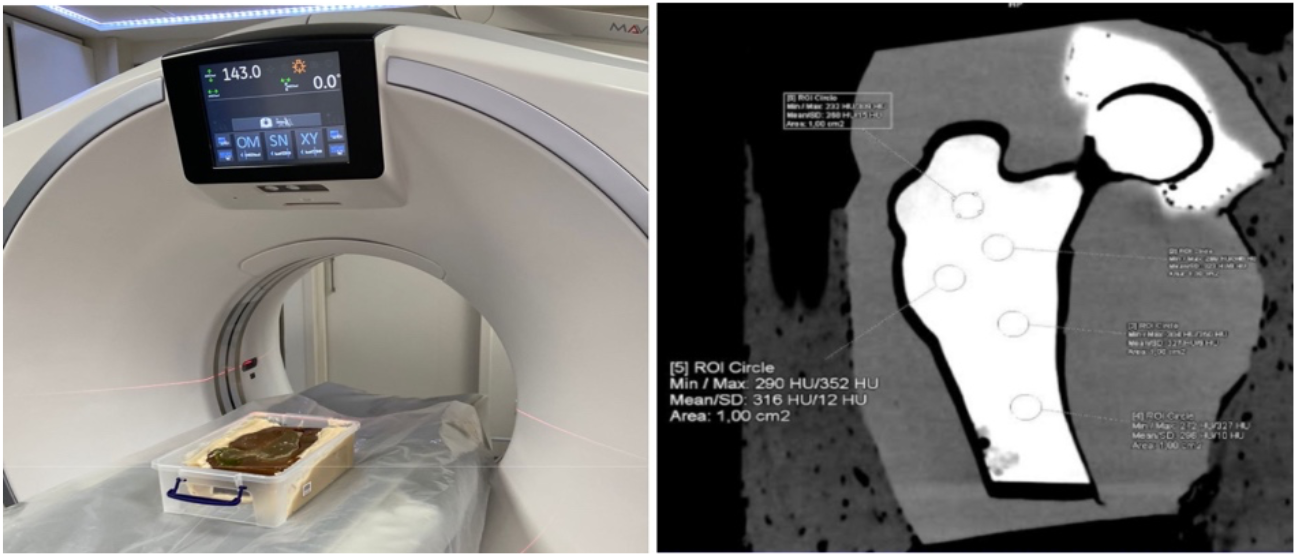
CT of the final phantom with the respective measurement of Hounsfield Units (HU). Final phantom in the CT scan (left image). The obtained image with the respective ROIs (right image).

## RESULTS

A total of 34 CT examinations of female patients with a bony pelvis protocol were selected, using a coronal slice with a soft tissue algorithm. The measurement of the different ROIs resulted in 442 data points, from which a 95% confidence interval was obtained for the femoral head, femoral neck, femoral cortex, femoral diaphysis, medial acetabulum, lateral acetabulum, muscle, and fat.

After obtaining the confidence intervals from the patient data, five material tests were conducted under CT, and ROIs were measured in each, resulting in 51 HU data points from the selected mixtures. The HU values of those mixtures that fell within any of the patient HU confidence intervals were selected, as shown in Table 1.

**Table 1:**
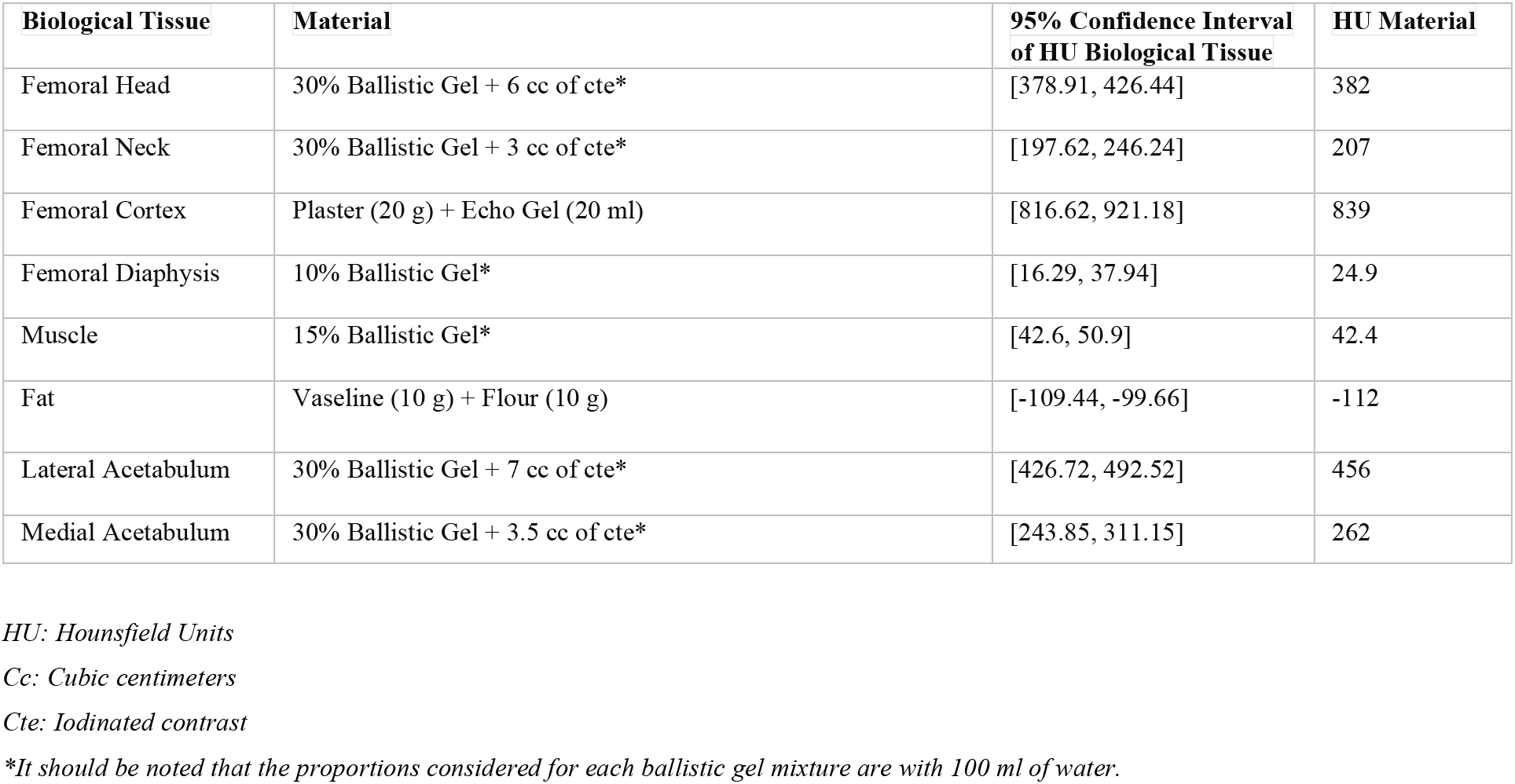
Material Corresponding to Each Biological Tissue of the Hip According to Patient Confidence Intervals and HU of the Materials.

It is observed that the HU values of the materials fall within the confidence intervals of the HU values of human tissues, with the exception of fat, which has a very close value.

For the construction of the phantom, it was decided to make the entire femur (head, neck, cortex, and diaphysis) and the entire acetabulum, both medial and lateral, using a single material for both structures: 30% ballistic gel + 6 cc of iodinated contrast.

The fabrication of the phantom commenced with the 3D printing of the hip and acetabulum in a positive form, followed by the creation of molds using RTV silicone rubber XL 8820T. Once the molds were prepared, we proceeded with the mixing stage, adhering to the corresponding proportions outlined in Table 1, taking into account material changes in both bone tissues while maintaining the materials for muscle and adipose tissues as specified in Table 1.

Stage III spanned a total of 4 days. The 3D printing of the femur and acetabulum required 12 hours (Figure 6), while the fabrication of silicone molds took 24 hours to dry and solidify. Subsequently, the mixing and assembly of the phantom were completed within 1 day. The crafting of acrylic boxes for the silicone mold and the creation of the 3D model of the hip each took 1 day.

**Figure 6.**
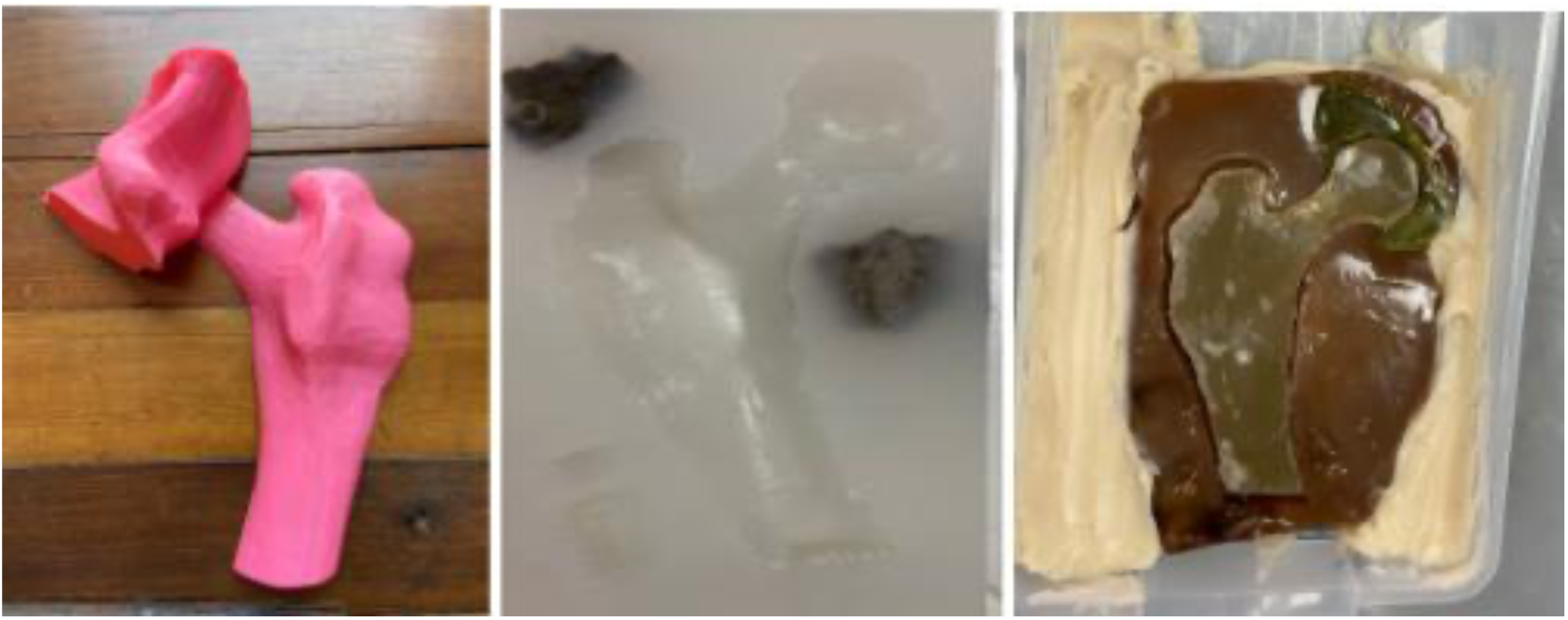
Construction of the final phantom. 3D Printing of the femur and acetabulum in positive form using PLA thermoplastic filament (right image). Negative silicone rubber RTV XL 8820 mold of the femur (center image). Right hip phantom with different tissues, bone, muscle, and adipose (left image).

Finally, the phantom test was conducted on the 64-channel General Electric CT scanner, using a bone pelvis protocol in soft tissue algorithm, similar to the tests conducted for the previous materials. The Hounsfield Units (HU) were measured in the different materials of the phantom using a 1 cm^2 Region of Interest (ROI), obtaining 28 data points for the phantom femur, 15 data points for the phantom acetabulum, 98 data points for phantom muscle, and 39 data points for phantom adipose tissue. These data were compared with the HU data from patients, where we have 136 data points for patient femurs, 102 data points for patient acetabula, 102 data points for patient muscles, and 102 data points for patient adipose tissue.

The HU values of human tissues and materials were compared in the following graph.

**Graphic 1.**
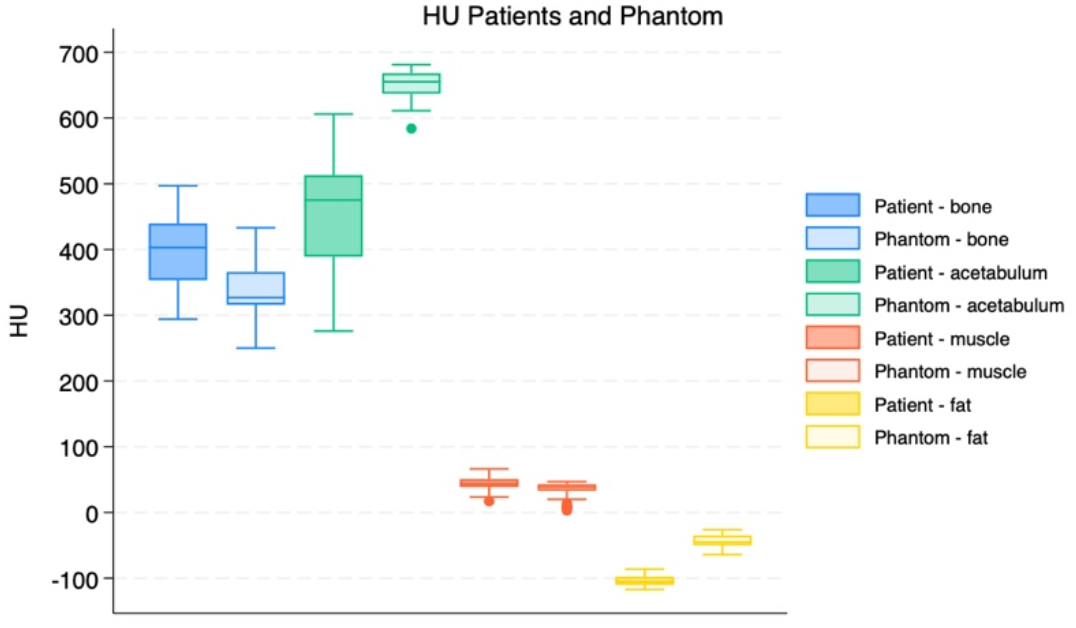
Comparison of Hounsfield Units (HU) between human tissues and selected materials in the final phantom. Figure 1. Boxplot of the Hounsfield Units (HU) for the different hip tissues of the patients and the HU for the different materials of the phantom. Significant similarity in values is observed across tissues, except for the acetabulum and adipose tissue, with the latter being the only tissue outside the confidence interval of human tissue.

## COSTS

To produce the final phantom, considering the amount of material used proportionally for the hip phantom, the prices of each material individually and the total cost of the phantom were calculated, resulting in $56,964 (Chilean pesos).

**Table 6:** Price of Materials Used in the Final Phantom.

| Material | Total Quantity | Price (\$) |
| --- | --- | --- |
| Gelatin | 486 g of gelatin | 4.860 |
| White Vinegar | 110 ml | 154 |
| Glycerin Soap | 110 ml | 203 |
| Iodinated Contrast Medium | 18 ml | 14.400 |
| Vaseline | 990 g | 8.415 |
| Flour | 660 g | 304 |
| Thermoplastic Filament (PLA) | 83 g | 1.328 |
| RTV XL Silicone Rubber 8820T | 1 kg | 22.000 |
| Plastic Box | 1 unit | 5.300 |
| **TOTAL** | | **\$56.964** |

## CONCLUSION AND DISCUSSION

The primary objective of designing an anthropomorphic phantom with materials resembling human tissues was achieved. Both bone and muscle tissues fell within the confidence interval (CI) estimated from the sample. However, the adipose tissue showed a HU value of -112, slightly higher than the estimated CI of -109.44 to -99.66. This CI was also slightly elevated compared to the estimates provided by Woodard & White (1986)(12), but it is quite close to the values reported in the literature. The muscle (15% ballistic gel) showed a value of 42.4 HU, which is within the CI for human tissue [42.6 to 50.9], and aligns with the values reported by Woodard & White, which ranged between 40 and 44 HU. Other authors, such as Sirtoli et al. (2017) (7), have reported slightly higher values of 60 HU, but with a margin of ± 30, indicating a broad range of HU values, thus resembling the findings of this study. Regarding the bone, the HU value of the material, which was 30% ballistic gel with 6 cc of contrast agent, was 382 HU, falling within the CI for human tissue [378.9 to 426.44]. This value is higher than the reported literature value for trabecular bone, which is 262 HU according to Mohammed et al. (2023)(20). However, it should be noted that the phantom was not constructed to distinguish between cortical and trabecular bone, but rather to simulate the femoral head. If we had detailed the construction for each specific bone structure (trabecular bone, cortical bone, and compact bone), all these would likely fall within the values reported in the literature and our human tissue sample. Given that this is a low-cost prototype made relatively quickly, we believe that in a subsequent stage, we can manufacture the phantom with greater precision.

The main disadvantages of this project are primarily the durability of the materials, which began to decompose quickly. The calculation of physical and radiological properties was not conducted as thoroughly as in the study by Mohammed et al. (2018)(14), and the design with aesthetic characteristics was not well achieved. However, as a prototype, it is undoubtedly improvable.

The primary advantages of this phantom are its low cost (56,000 Chilean pesos), which is very similar to, or even lower than, the cost reported by Mohammed et al. (2023) (20) (53 British pounds). The methodology, cost, and similarity to human tissue are the great advantages of this phantom. It can aid radiology services in more accurately estimating effective doses and contribute to the education of radiology and anatomy students. With this methodology, phantoms can be created to simulate various pathologies and scenarios, offering unlimited possibilities. We will continue to refine our models to make them more precise and aesthetically pleasing.

## Data Availability

All data produced in the present study are available upon reasonable request to the authors

